# Inflammatory Biomarkers and Interpretable Machine Learning Model for Stroke-Associated Pneumonia Risk Stratification in Patients Undergoing Bridging Therapy for Acute Ischemic Stroke

**DOI:** 10.64898/2026.04.15.26350997

**Authors:** Xue-Yao Wang, Miao-Miao Li, Shuang-Mo Zhao, Xiao-Yu Jia, Wen-Song Yang, Lu-Lu Chang, Hui-Min Wang, Jian-Ting Zhao

**Author notes:** Xue-Yao Wang and Miao-Miao Li share first-author status and contributed equally to this paper. **Correspondence:** Hui-Min Wang or Jian-Ting Zhao or.

## Abstract

Stroke-associated pneumonia (SAP) is a major complication affecting the prognosis of acute ischemic stroke (AIS) patients who undergo intravenous thrombolysis followed by bridging mechanical thrombectomy (MT). Existing predictive tools are mostly subjective, and the value of combined biomarkers and interpretable machine learning (ML) models in this specific population remains unclear. This study aimed to construct an interpretable ML model to predict SAP risk in AIS patients undergoing bridging therapy, and to identify key predictive factors using Shapley Additive Explanations (SHAP). A single-center retrospective observational study was conducted on 135 AIS patients who received intravenous thrombolysis followed by bridging MT at Xinxiang Central Hospital from January 2019 to December 2023. Clinical data, laboratory indicators (including neutrophil-to-lymphocyte ratio [NLR], platelet-to-lymphocyte ratio [PLR], systemic immune-inflammation index [SII], and systemic inflammatory response index [SIRI]) were collected. LASSO regression was used for feature selection, and ten ML models were constructed to predict SAP. The optimal model was selected based on area under the receiver operating characteristic curve (AUC-ROC) and decision curve analysis (DCA), and SHAP analysis was applied to interpret the model. Among 135 included patients, 70 (51.9%) developed SAP. LASSO regression selected 11 key variables associated with SAP. The CatBoost model showed the best performance, with an AUC of 0.952 in the training set and 0.932 in the test set. SHAP analysis revealed that the 7-day National Institutes of Health Stroke Scale (NIHSS_7d), 24-h SIRI (SIRI_24h), and 24-h white blood cell count (WBC_24h) were the top three factors contributing to SAP prediction. Dynamic detection showed that 24-h and 48-h NLR, 24-h SII, and 24-h and 48-h SIRI were significantly higher in the SAP group than in the non-SAP group (all P < 0.05), while no significant difference in PLR was observed between the two groups. Inflammatory biomarkers (24-h NLR, 24-h SII, 24-h SIRI, 48-h NLR, 48-h SIRI) are closely associated with SAP in AIS patients undergoing bridging therapy. The interpretable CatBoost model constructed with LASSO-selected variables exhibits high predictive value for SAP, which can help clinicians identify high-risk patients early and guide personalized treatment.

## 1 Introduction

Acute ischemic stroke is the most common type of stroke in China, accounting for approximately 69.6%–72.8% of cases of stroke (1). Its high rates of disability and mortality make it the second leading cause of death and the third leading cause of disability, respectively. Among these, stroke-associated infections (SAIs) are important factors affecting prognosis and mortality. SAIs are defined as infections occurring within 7 days after stroke, primarily including pneumonia and urinary tract infections, with pneumonia particularly linked to poorer clinical outcomes(2, 3). Endovascular therapy (EVT) can significantly improve mortality and disability rates in patients with acute ischemic stroke due to large-vessel occlusion; however, postoperative infections may still affect recovery, and the efficacy of prophylactic antibiotics has not met expectations(4). Widespread inappropriate antibiotic use may exacerbate the risk of SAIs or lead to antibiotic resistance (5). Therefore, stratifying patients who may benefit from antibiotics through biomarker screening may help improve prognosis.

Various risk factors for stroke-associated pneumonia (SAP) have been identified, including sex, age, and stroke severity (6, 7). Existing clinical prediction models, such as the Adapted Symptom Distress Scale (A2DS2) and acute ischemic stroke-associated pneumonia score (AIS-APS), are used for early SAP prediction; however, they mostly rely on clinical symptoms, which are subjective and may delay early identification (6, 8). In contrast, Hasse et al. indicated that the use of combined biomarkers has predictive value for SAI (9), highlighting the potential of objective laboratory indicators to improve prediction accuracy. Thus, further improvements are required to develop simple and accessible predictive tools based on biomarkers. Autonomic nervous system activation due to ischemic stroke leads to the release of pro-inflammatory mediators, resulting in long-term immune suppression and an increased risk of infection (10). When brain tissue is damaged, immune cells release cytokines, leading to systemic immune activation and suppression (11). This sustained inflammatory response ultimately depletes the immune system, leading to lymphocyte reduction and further increasing the incidence of post-stroke infections (12, 13). Biomarkers such as the neutrophil-to-lymphocyte ratio (NLR), platelet-to-lymphocyte ratio (PLR), systemic immune-inflammation index (SII), and systemic inflammatory response index (SIRI) are associated with inflammation and immune status and may predict the occurrence of pneumonia after acute ischemic stroke. NLR is an important indicator for assessing systemic inflammation and infection risk and can predict community-acquired pneumonia and outcomes in patients with ischemic stroke(14, 15). In contrast, PLR is a marker based on platelet aggregation and systemic inflammation (16). PLR may be an independent risk factor for SAP in patients with stroke (17). The SII and SIRI indicate a balance between the inflammatory response and immune status and have good predictive capability for SAP in patients with acute ischemic stroke (18, 19). Previous studies indicated an association between these biomarkers and SAP; however, these studies often fail to capture the complex interactions between multiple biomarkers and clinical factors, limiting the accuracy of prediction. Machine learning (ML) is a computational algorithm that learns from data and identifies underlying patterns, capable of uncovering complex relationships within large datasets, making it of significant clinical value for improving the prediction accuracy of SAP (20, 21). Rooted in cooperative game theory, Shapley Additive Explanations (SHAP) quantifies the contribution of each input feature to individual predictions, effectively addressing the “black-box” limitation of complex machine learning models. To date, no studies have incorporated SHAP into machine learning (ML) models for predicting stroke-associated pneumonia (SAP) in acute ischemic stroke patients who underwent intravenous thrombolysis followed by bridging mechanical thrombectomy. The model developed in this study can accurately predict SAP risk in this specific population, enabling clinicians to identify high-risk individuals early and guide personalized treatment decisions.

## Materials and methods

### 1.1 Patients and participants

This single-center retrospective observational study was conducted at Xinxiang Central Hospital. Patients with acute ischemic stroke admitted continuously from January 1, 2019 to December 31, 2023 who underwent bridging MT after intravenous thrombolysis were enrolled. The study was conducted in accordance with the guidelines of the 1964 Declaration of Helsinki and its later amendments. Ethical approval was obtained from the Medical Ethics Committee of Xinxiang Central Hospital (Approval No.: 2025-153-01 (K))

Patients who met the following criteria were included: (i) aged >18 years with acute ischemic stroke, symptom onset within 4.5 h, diagnosis consistent with the “Chinese Guidelines for the Diagnosis and Treatment of Acute Ischemic Stroke 2023” (22), and confirmed by imaging; (ii) underwent intravenous thrombolysis followed by endovascular MT, consistent with the “Chinese Guidelines for Endovascular Treatment of Acute Ischemic Stroke 2023” (23).

Patients were excluded based on the following criteria: absence of routine blood indicators at admission and 24 and 48 h, active infection within 2 weeks before admission, infectious diseases, autoimmune disorders requiring immunosuppressive treatment, malignant tumors, blood disorders, severe liver disease, kidney disease, and significant trauma or surgery(16, 24).

### 1.2 Clinical assessment

Data collected from medical records included demographic characteristics, laboratory data, medical history, clinical features, and imaging information such as sex, age, history of hypertension, diabetes, chronic obstructive pulmonary disease, current smoking and drinking habits, fasting blood glucose, total cholesterol, C-reactive protein, time windows for thrombolysis and surgery, scores from the National Institutes of Health Stroke Scale (NIHSS, where higher scores indicate worse neurological function), modified Rankin Scale (mRS), and modified treatment in cerebral infarction grading. Laboratory tests for complete blood count and other laboratory indicators such as total cholesterol and C-reactive protein were performed at admission, 24 h, and 48 h using a fully automated biochemical analyzer (AU2700; Olympus, Tokyo, Japan), following the manufacturer’s guidelines. The NLR, PLR, SII, and SIRI were calculated using the following formulas: NLR = neutrophil count / lymphocyte count; PLR = platelet count / lymphocyte count; SII = (neutrophil count × platelet count) / lymphocyte count; SIRI = (neutrophil count × monocyte count) / lymphocyte count.

A total of 192 patients were initially screened for eligibility based on the inclusion criteria. Among these, 57 were excluded according to the exclusion criteria, resulting in 135 patients included in the final analysis (Figure 1). The patients were grouped based on the presence or absence of pneumonia (pneumonia or non-pneumonia groups, Figure 1). The diagnostic criteria for SAP were based on the guidelines for stroke-associated pneumonia (25), which include new or progressive patchy infiltrates, solid lung lobe changes, or pleural effusion on chest X-ray or computed tomography, along with two or more of the following clinical symptoms: fever (temperature >38℃), purulent respiratory secretions, and abnormal white blood cell count (either <4000 × 10⁹/L or >10000 × 10⁹/L).

**Figure 1.**
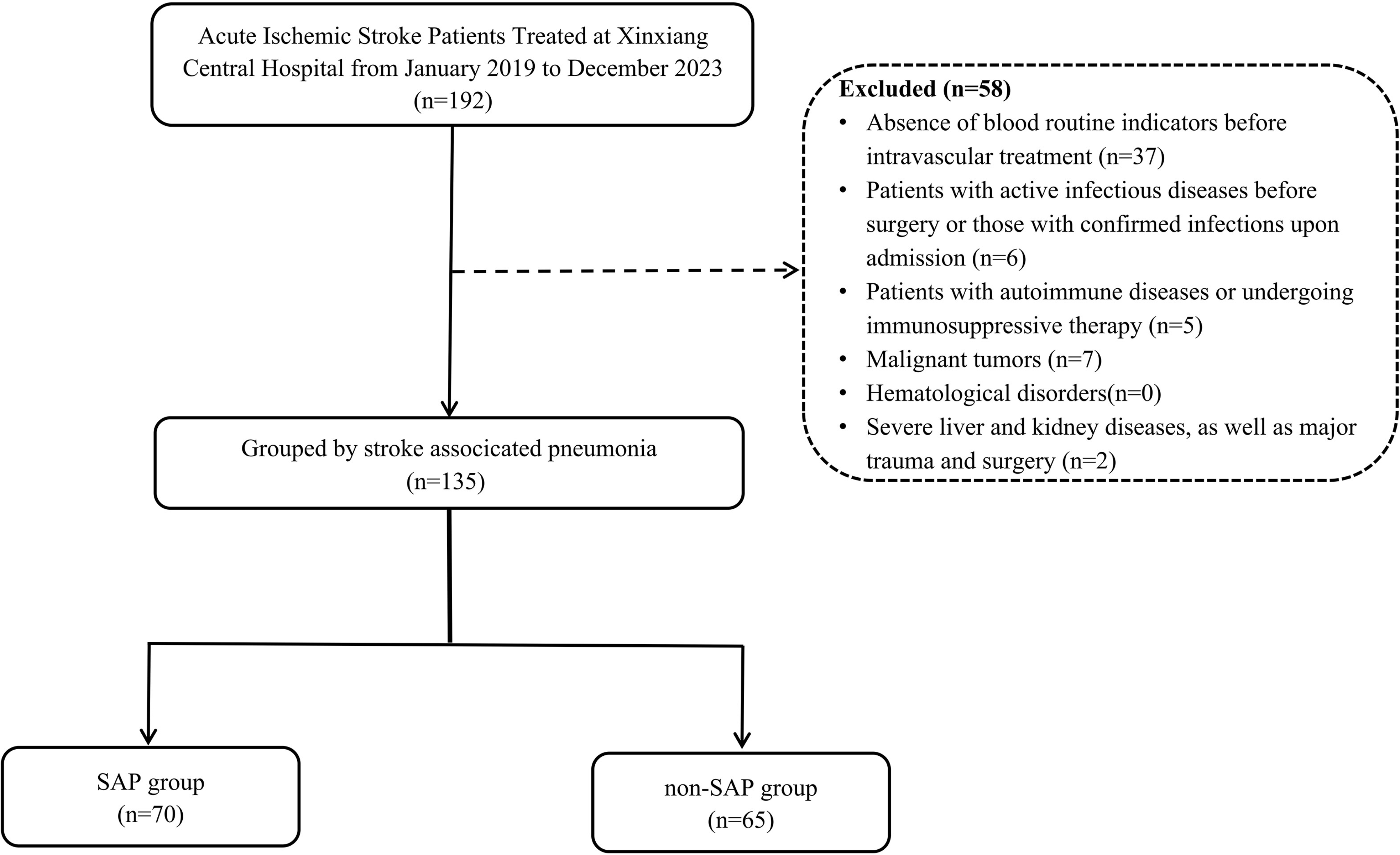
The flow chart of excluded patients.

### 1.3 Statistical analysis

Continuous variables are presented as mean ± standard deviation or median (interquartile range). Normally distributed and non-normally distributed data were assessed using t-tests and the Mann–Whitney U test, respectively. Categorical variables are expressed as numbers with percentages and compared using the chi-squared or Fisher’s exact tests, as appropriate.

The dataset was randomly split into a training cohort (70%, n=95) and a validation cohort (30%, n=40) using a stratified random sampling method based on SAP status to ensure balanced distribution of SAP cases between the two cohorts. To reduce dimensionality and select the most predictive features, LASSO regression was employed to screen variables that were significantly associated with SAP in univariate analysis (P < 0.05). Additionally, variance inflation factor (VIF) was used to assess and mitigate multicollinearity among the selected variables (VIF < 10 was considered no significant multicollinearity). As the penalty term (λ) increased, the regression coefficients of the model’s predictors were progressively shrunk toward zero. Ultimately, eleven variables with non-zero coefficients were selected, including: Age, NIHSS_7d, WBC_24h, SII_24h, SIRI_24h, NLR_48h, FBG, D-dimer, G-tube, NIHSS_preMT, NIHSS_24h_postIVT. Ten machine learning (ML) models were used to predict the risk of SAP: Random Forest (RF), Logistic Regression (LR), Gradient Boosting Machine (GBM), Neural Network (NN), Extreme Gradient Boosting(XGBoost), K-Nearest Neighbors (KNN), Adaptive Boosting (AdaBoost), Light Gradient Boosting Machine(LightGBM), Category Boosting (CatBoost), and Support Vector Machine (SVM). To assess model performance, Model evaluation was performed using the area under the receiver operating characteristic curve (AUC-ROC) and decision curve analysis (DCA). The model’s efficiency and effectiveness in managing stroke-associated pneumonia (SAP) in patients with acute ischemic stroke were further evaluated on both the test and validation sets using metrics including Accuracy, Precision, Recall, and F1-Score. SHAP interpretability analysis To enhance the interpretability of our model, we employed the SHAP method. SHAP is based on the concept of Shapley values from cooperative game theory, and it uses additive models to assess the contribution of each feature to the model’s predictions. This method provides a deeper understanding of model outputs, making it particularly useful for explaining complex ML models. We utilized the shapviz package to calculate and visualize the SHAP values of the CatBoost model, thereby identifying the key factors associated with SAP in acute ischemic stroke patients. In this study, we followed the SHAP guidelines (https://github.com/slundberg/shap) to interpret and visualize the CatBoost model. The SHAP histograms, SHAP bee swarm plots, and waterfall plots were generated. A higher absolute value of SHAP indicates a greater influence of the feature on the prediction outcome. Statistical analyses were performed using SPSS.26.0 (IBM, Armonk, NY, USA), GraphPad Prism.9.0 (GraphPad Software, San Diego, CA, USA), and R.4.4.1 (R Foundation for Statistical Computing, Vienna, Austria).

## 2 Results

In total, 135 patients with acute ischemic stroke who underwent intravenous thrombolysis and bridging MT were included in the study, of whom 70 (51.9%) developed SAP. The average age of the SAP group was 67.21 ± 12.21 years, including 26 females (37.1%). The average length of hospital stay was 15.5 days (9.75 and 24), with 51 cases of LAA (72.9%) and 38 cases of dysphagia (54.3%). The NIHSS score before thrombolysis was 19.5 (15 and 27). The baseline characteristics of patients with acute ischemic stroke who underwent intravenous thrombolysis and bridging MT are presented in Table 1. Patients with SAP exhibited higher NIHSS scores at 24 h post-thrombolysis (P = 0.002), higher pre-operative NIHSS scores (P = 0.011), higher NIHSS scores at 24 h post-surgery (P = 0.004), and higher NIHSS scores at 2–7 days post-admission (P < 0.001). The mRS score at discharge was also significantly higher (P < 0.001). The 90-day mRS score was also significantly higher in the SAP group (P < 0.001). These results suggest that poor neurological function is associated with an increased risk of stroke-related pneumonia and poor short-term outcomes. Higher fasting blood glucose levels were observed at admission (P = 0.010), along with elevated D-dimer levels (P = 0.012) and increased fibrin degradation products (P = 0.003), suggesting that hyperglycemia and a hypercoagulable state may promote immune-inflammatory responses, leading to the development of pneumonia. In the SAP group, neutrophil count at 24 h (P < 0.001), white blood cell count at 24 h (P = 0.002), neutrophil count at 48 h (P = 0.027), 24-h NLR (P = 0.001), 24-h SII (P = 0.025), 24-h SIRI (P < 0.001), 48-h NLR (P = 0.007), and 48-h SIRI (P = 0.024) were all significantly higher compared to the non-SAP group (Table 2).

**TABLE 1.**
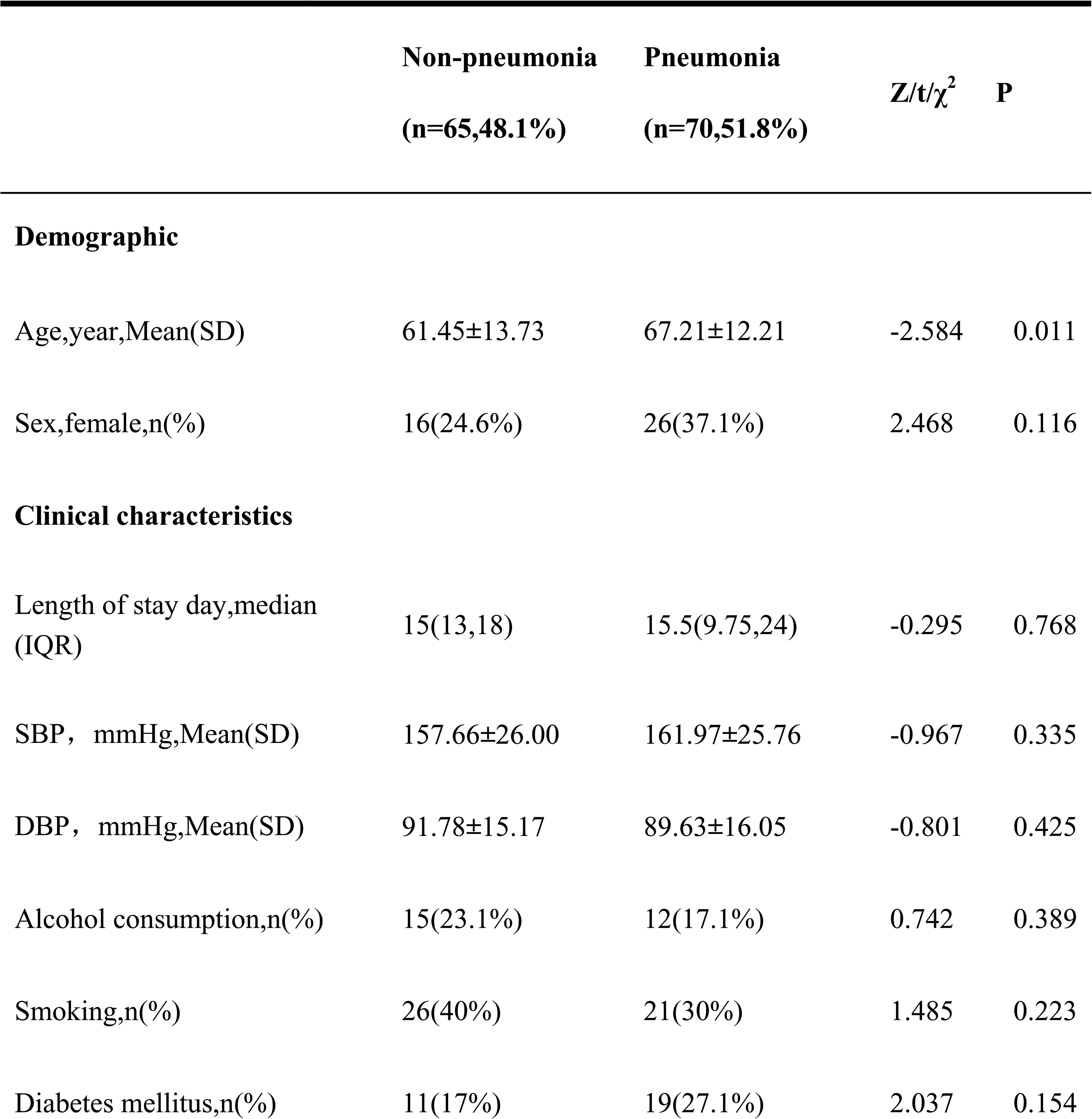
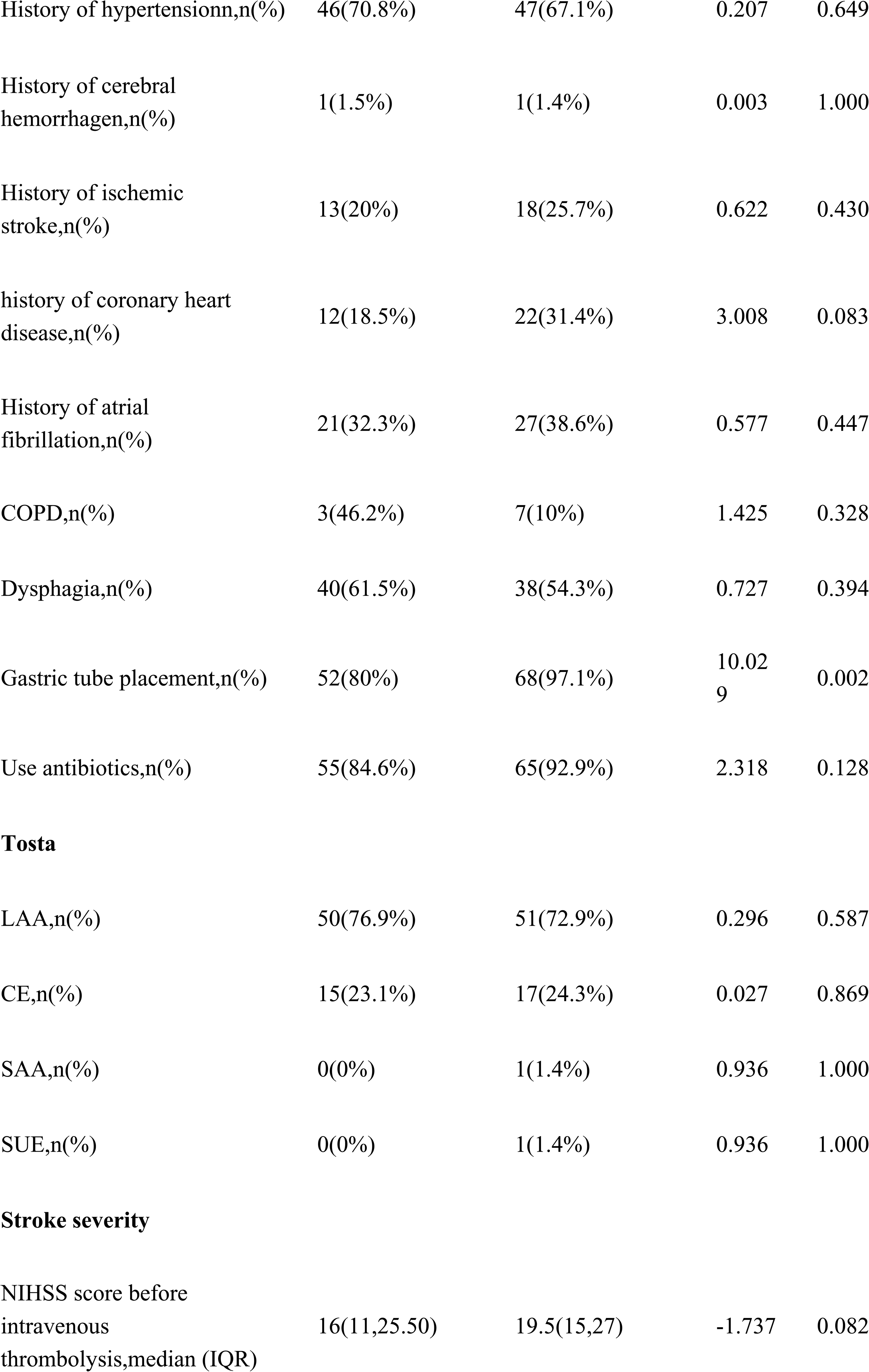
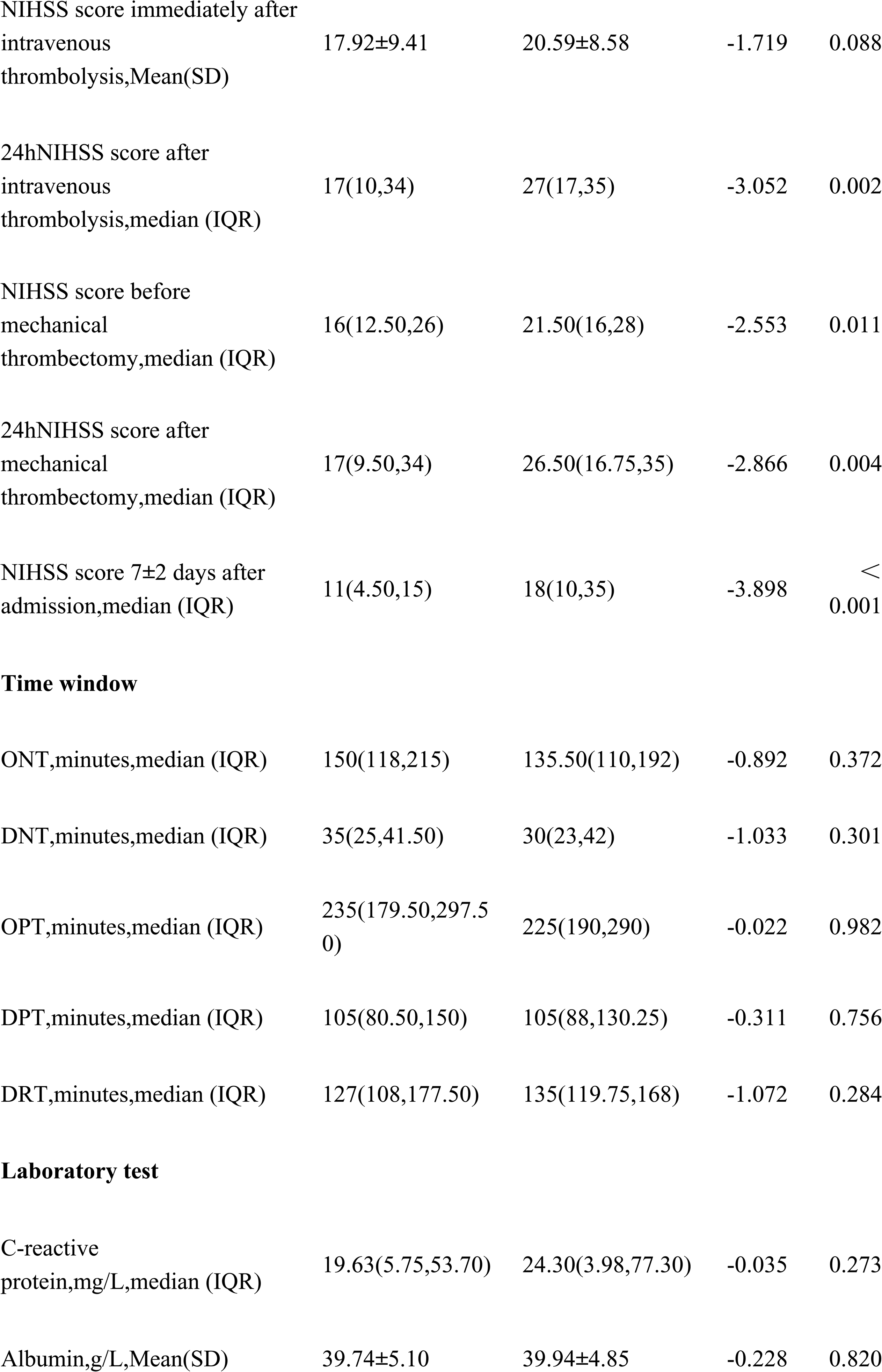
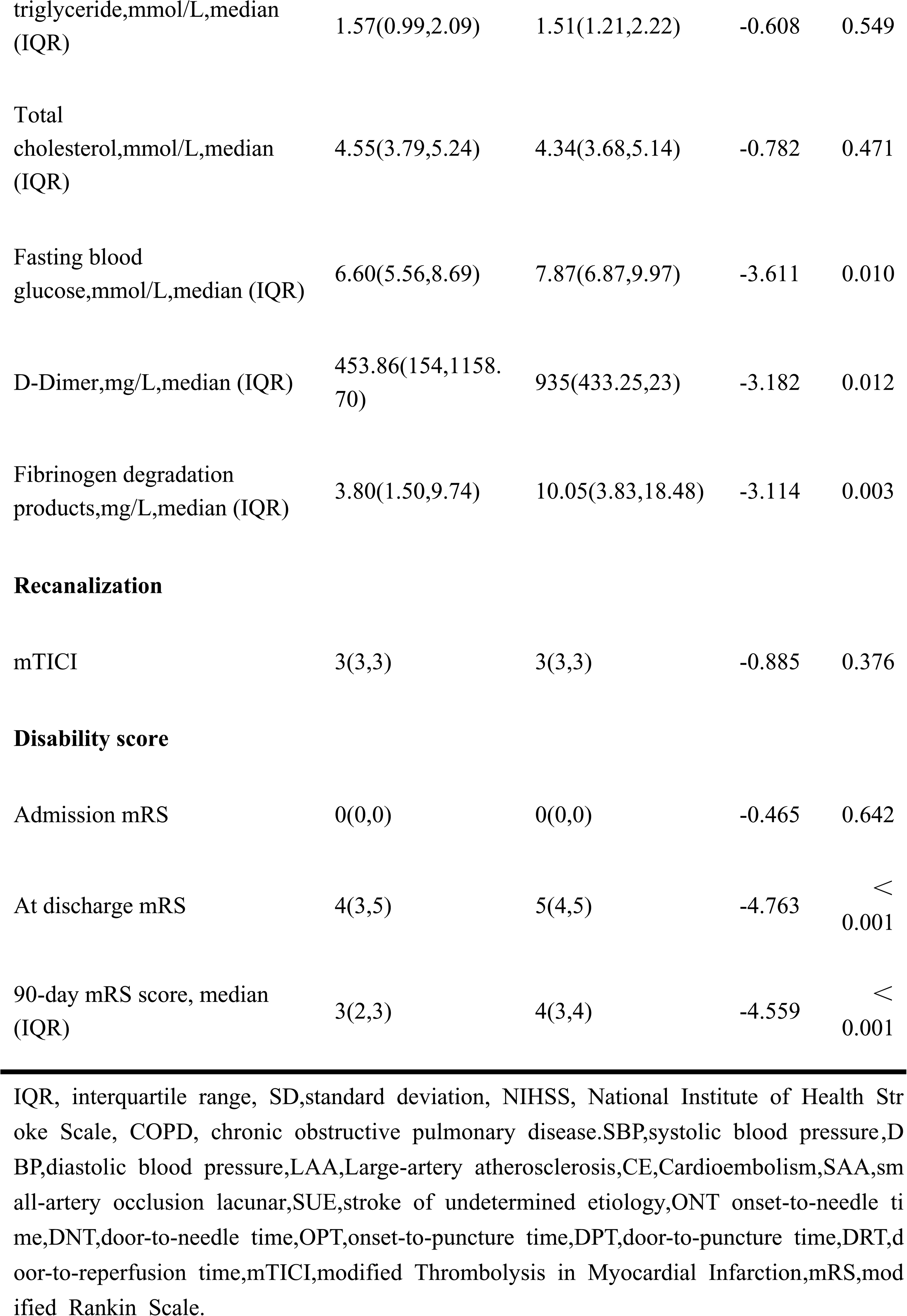
Baseline characteristics of the 135 patients with Intravenous thrombolytic bridging mechanical thrombectomy.

**TABLE 2.**
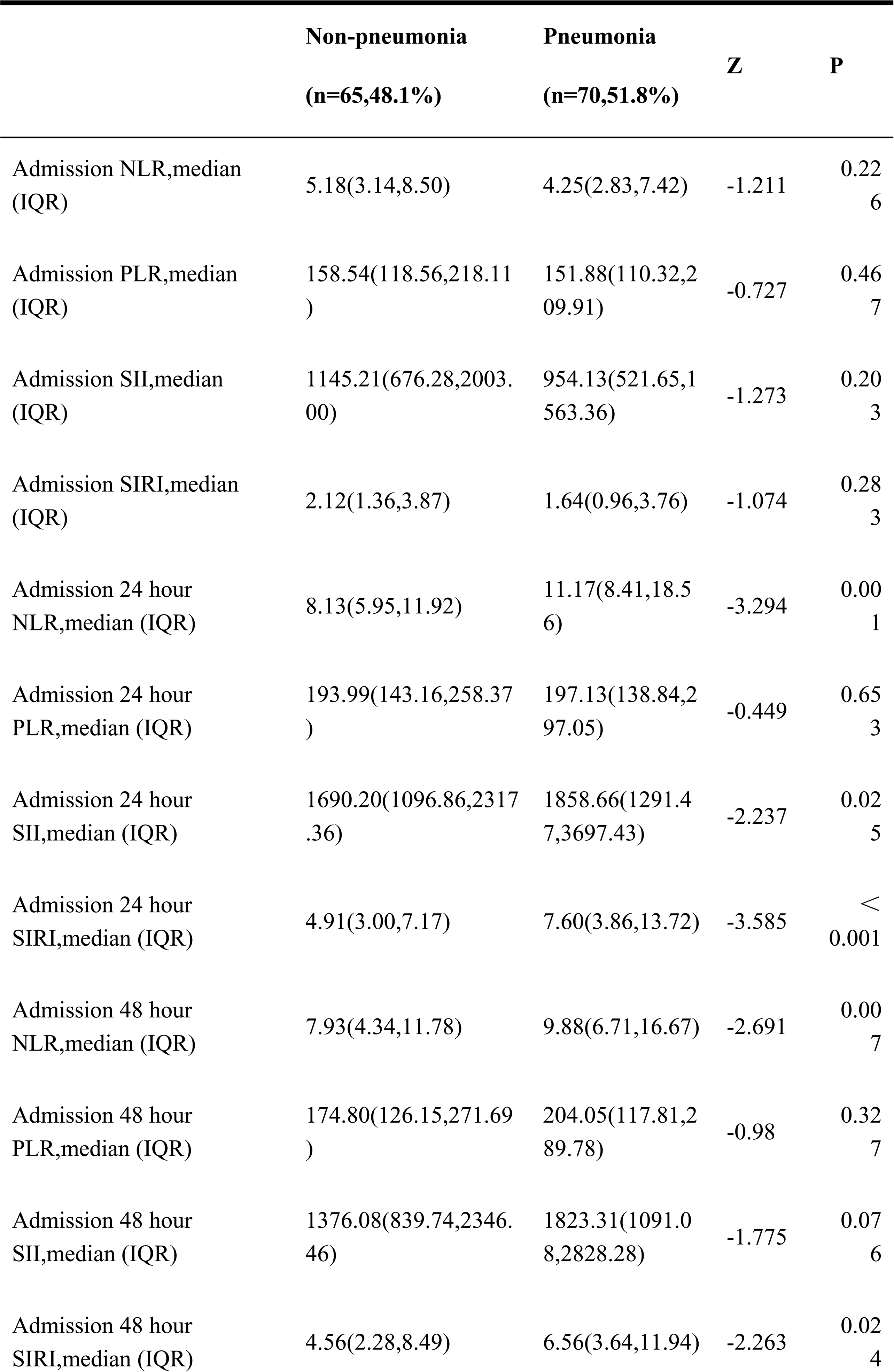
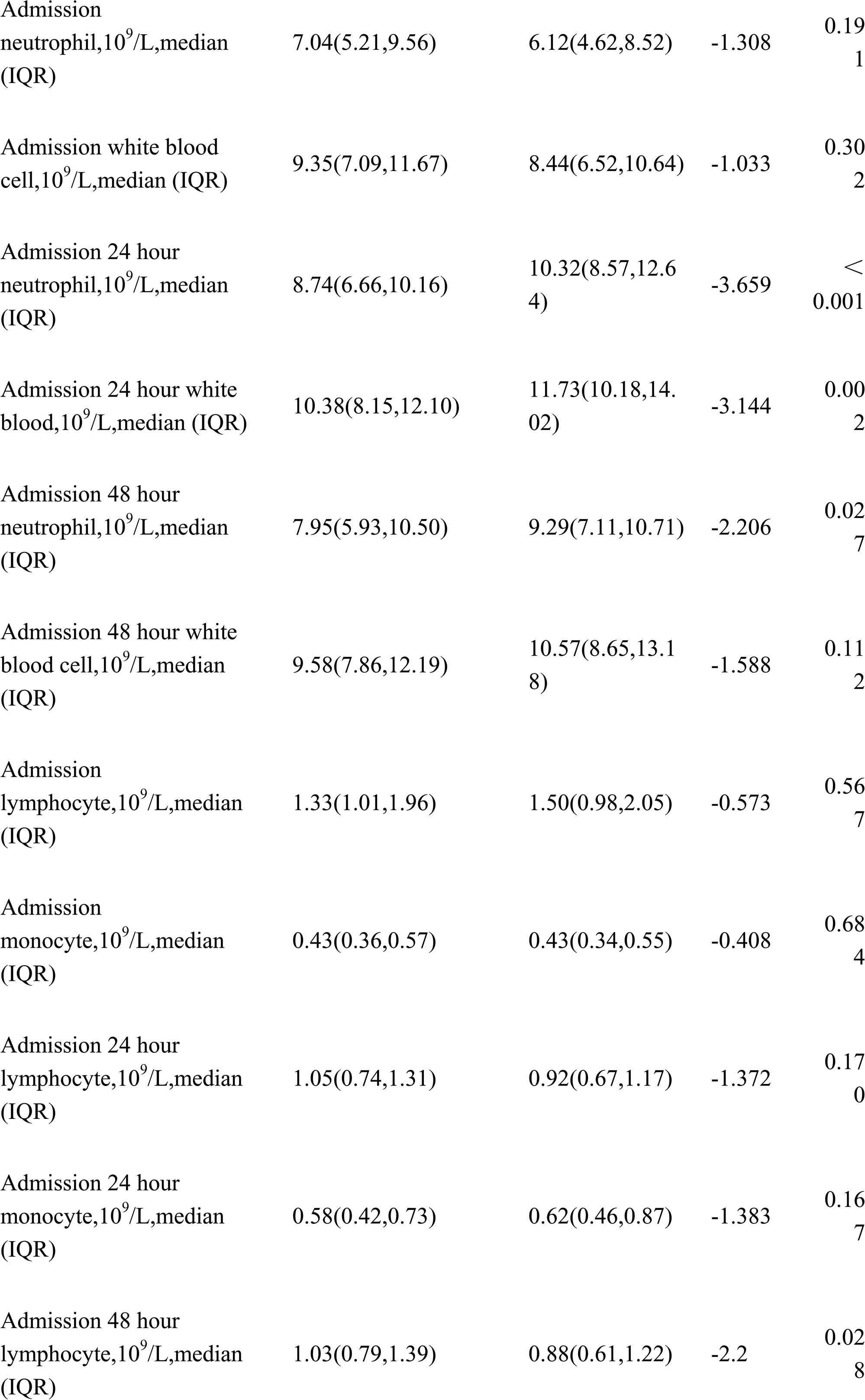
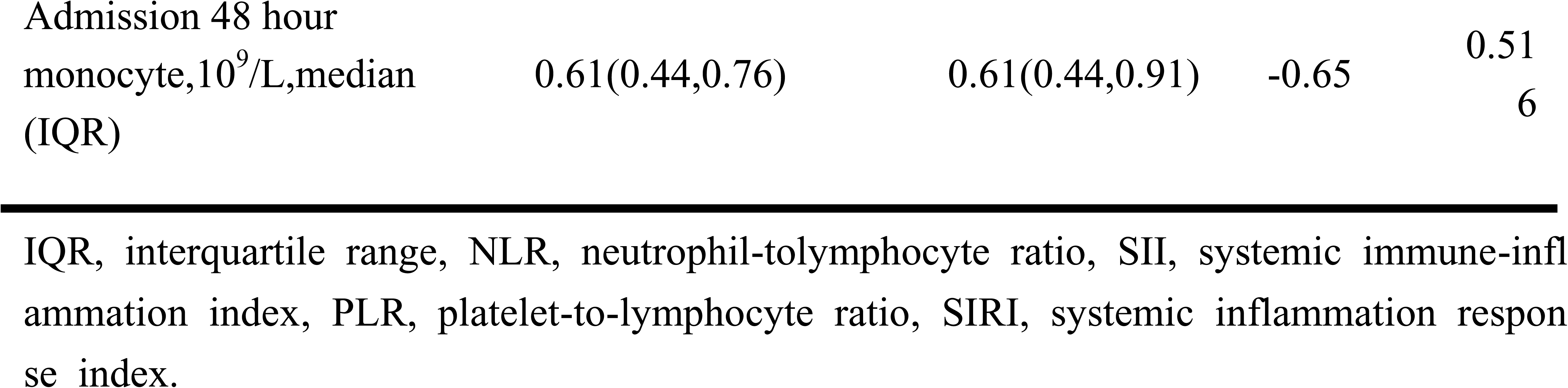
Inflammatory index of the 135 patients with Intravenous thrombolytic bridging mechanical thrombectomy.

In the analysis of the temporal changes in these biomarkers, significant differences in NLR levels between the SAP and non-SAP groups were observed at 24 and 48 h (24-h NLR, P < 0.01; 48-h NLR, P < 0.01), as illustrated in Supplemental Figure 1A. No differences were observed in the PLR between the two groups at admission, 24 h, or 48 h (Supplemental Figure 1B).

Significant differences were also observed in 24-h SII (24-h SII P < 0.01) (Supplemental Figure 1C), as well as in 24-h and 48-h SIRI (24-h SIRI P < 0.001, 48-h SIRI P < 0.05) (Supplemental Figure 1D).

In the temporal analysis of biomarkers, the SAP group showed significant increases in NLR, PLR, SII, and SIRI from admission to 24 h (Supplemental Figure 1A).

Using LASSO regression, we selected 11 important variables from 63 clinical features associated with the occurrence of SAP (Supplemental Figure 2 and Supplemental Figure 3). Based on these variables, we developed 10 ML models for prediction. After excluding overfitted models, we performed a comprehensive evaluation using ROC curves and DCA. The CatBoost model demonstrated the best performance, with an AUC of 0.952 on the training set and 0.932 on the test set, significantly outperforming the other models. Therefore, CatBoost was selected as the final predictive model (Figure 2, Figure 3, Table 3 and Table 4). In addition, this study employed additional performance metrics to evaluate model effectiveness, as detailed in Tables 3 and 4.

**Figure 2.**
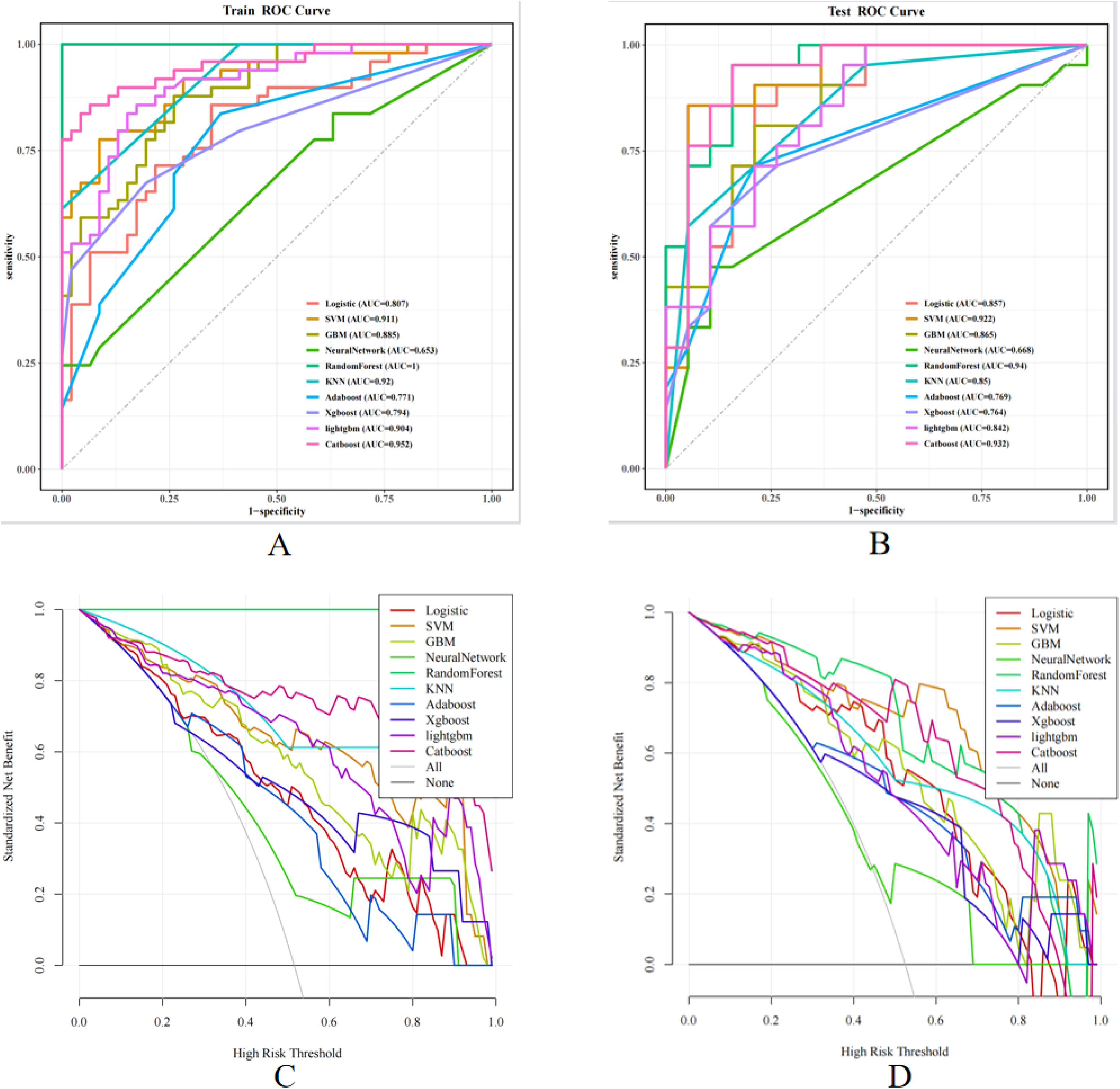
Model performance evaluation via ROC curves and decision curve analysis. (A) ROC curve of the training set; (B) ROC curve of the test set; (C) DCA curve of the training set; (D) DCA curve of the test set.

**Figure 3.**
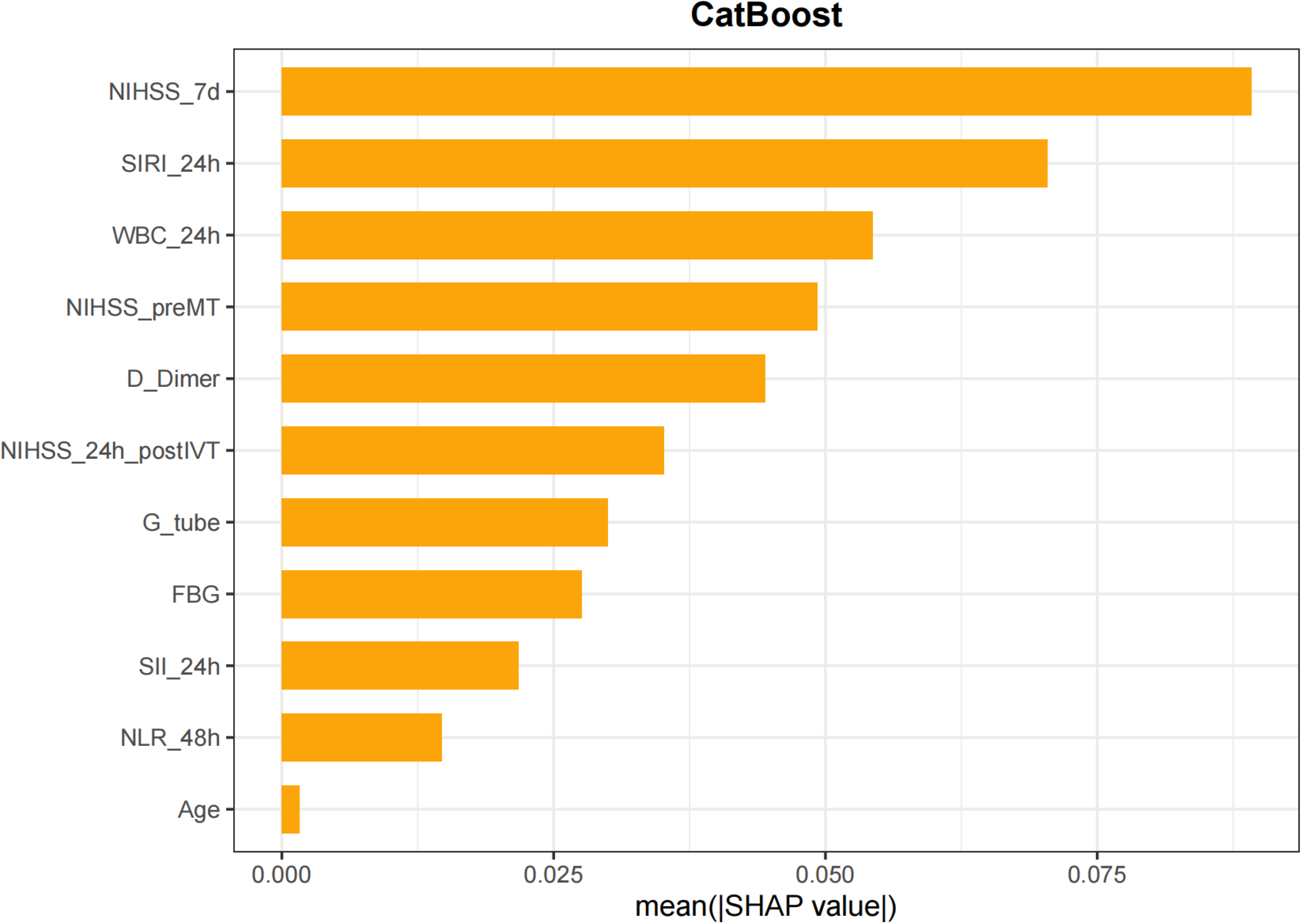
SHAP feature importance plot for the CatBoost model.

**TABLE 3.** Performance metrics of machine learning models on the training set.

| Model | Accuracy | Sensitivity | Specificity | Precision | F1 | AUC |
| --- | --- | --- | --- | --- | --- | --- |
| Logistic | 0.758 | 0.857 | 0.652 | 0.724 | 0.785 | 0.807 |
| SVM | 0.842 | 0.776 | 0.913 | 0.905 | 0.835 | 0.911 |
| GBM | 0.811 | 0.878 | 0.739 | 0.782 | 0.827 | 0.885 |
| NeuralNetwork | 0.611 | 0.245 | 1 | 1 | 0.393 | 0.653 |
| RandomForest | 1 | 1 | 1 | 1 | 1 | 1 |
| KNN | 0.8 | 0.612 | 1 | 1 | 0.759 | 0.92 |
| Adaboost | 0.737 | 0.837 | 0.63 | 0.707 | 0.766 | 0.771 |
| Xgboost | 0.737 | 0.673 | 0.804 | 0.786 | 0.725 | 0.794 |
| lightgbm | 0.842 | 0.837 | 0.848 | 0.854 | 0.845 | 0.904 |
| Catboost | 0.895 | 0.837 | 0.957 | 0.953 | 0.891 | 0.952 |

The table reports threshold, accuracy, sensitivity, specificity, precision, and F1-score for each ML model trained to predict pneumonia in AIS patients with IVT bridging MT. Logistic, Logistic Regression SVM, Support Vector Machine, GBM, Gradient Boosting Machine, KNN, K-Nearest Neighbor, Adaboost, Adaptive Boosting, Xgboost, Extreme Gradient Boosting, lightgbm, Light Gradient Boosting Machine, Catboost, Category Boosting

**TABLE 4.** Performance metrics of machine learning models on the test set.

| Model | Accuracy | Sensitivity | Specificity | Precision | F1 | AUC |
| --- | --- | --- | --- | --- | --- | --- |
| Logistic | 0.825 | 0.857 | 0.789 | 0.818 | 0.837 | 0.857 |
| SVM | 0.9 | 0.857 | 0.947 | 0.947 | 0.9 | 0.922 |
| GBM | 0.8 | 0.81 | 0.789 | 0.81 | 0.81 | 0.865 |
| NeuralNetwork | 0.675 | 0.476 | 0.895 | 0.833 | 0.606 | 0.668 |
| RandomForest | 0.9 | 0.952 | 0.842 | 0.87 | 0.909 | 0.940 |
| KNN | 0.75 | 0.571 | 0.947 | 0.923 | 0.706 | 0.850 |
| Adaboost | 0.75 | 0.714 | 0.789 | 0.789 | 0.75 | 0.769 |
| Xgboost | 0.725 | 0.571 | 0.895 | 0.857 | 0.686 | 0.764 |
| lightgbm | 0.775 | 0.952 | 0.579 | 0.714 | 0.816 | 0.842 |
| Catboost | 0.9 | 0.952 | 0.842 | 0.87 | 0.909 | 0.932 |

This table presents the performance of each ML model in predicting pneumonia among AIS patients with IVT bridging MT in the test set. Logistic, Logistic Regression SVM, Support Vector Machine, GBM, Gradient Boosting Machine, KNN, K-Nearest Neighbor, Adaboost, Adaptive Boosting, Xgboost, Extreme Gradient Boosting, lightgbm, Light Gradient Boosting Machine, Catboost, Category Boosting

SHAP analysis and model interpretability SHAP histograms and SHAP beeswarm plots were used to visualize the contributions of multiple features in the CatBoost model for predicting SAP. The plots revealed that NIHSS_7d showed a large positive SHAP value at higher levels, indicating a substantial contribution to the model’s prediction. Specifically, higher NIHSS_7d values were associated with an increased risk of SAP.

Additionally, higher values of SIRI_24h and WBC_24h also corresponded to stronger positive SHAP values, further supporting the model’s prediction of event occurrence. Conversely, features such as Age and NLR_48h had smaller SHAP values, suggesting their limited contribution to the model. Notably, when the values of these features were elevated, their SHAP values approached zero, indicating minimal impact on the prediction of SAP(Figure 4).

**Figure 4.**
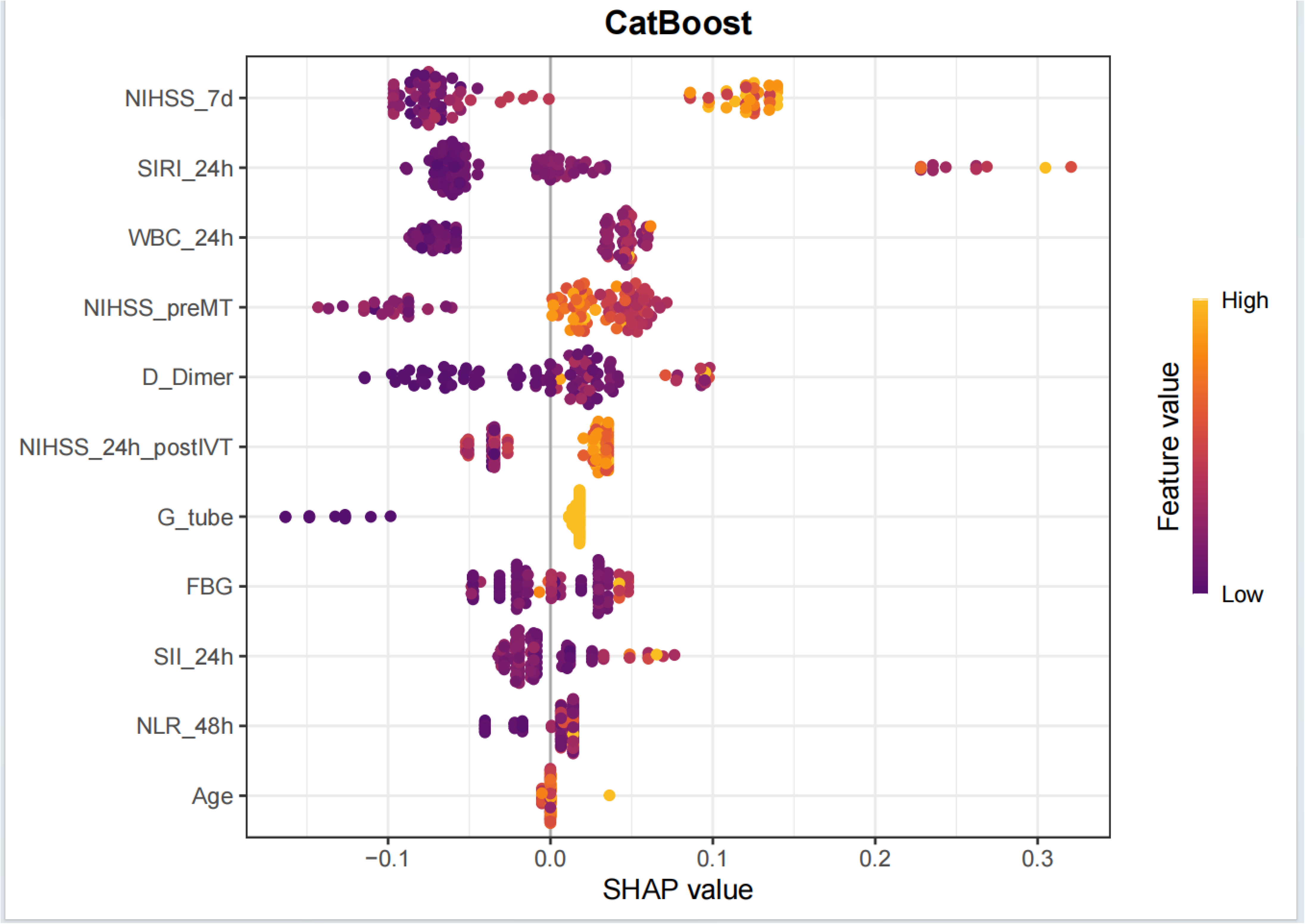
SHAP summary plot for the CatBoost prediction model.

We also demonstrated the interaction between different variables. The SHAP scatter plot visualized the interaction between Age and NIHSS_24h_postIVT in the CatBoost model’s prediction of SAP.

The plot showed that samples with most ages and low NIHSS_24h_postIVT corresponded to SHAP values near zero, indicating that these feature combinations contributed minimally to the model’s prediction, possibly reflecting a lower risk of SAP. In contrast, when NIHSS_24h_postIVT was elevated and age was advanced, the SHAP values increased notably, suggesting that this combination of features played a larger role in the model’s prediction, highlighting the association between high NIHSS_24h_postIVT and advanced age with the occurrence of SAP (Supplemental Figure 4).

The visualization results of SHAP waterfall plot and SHAP force plot demonstrates the performance of the CatBoost model in individual SAP prediction. For a patient who developed SAP, the SHAP value analysis revealed that several features contributed to the increased risk (e.g., NIHSS_preMT, SHAP value = 0.0542), while other features had negative SHAP values, exerting a protective effect. Ultimately, the combined effect of these pro-risk and protective features resulted in a predicted SAP risk of 0.326 for this patient, which may be attributed to the limited sample size or other unmeasured confounding factors. (Supplemental Figure 5 and Figure 6).

## 3 Discussion

This study employed LASSO regression combined with 10 common ML algorithms to construct a predictive model for identifying the risk of SAP in acute ischemic stroke patients undergoing bridging therapy. Additionally, the SHAP method was used to interpret and visualize the CatBoost model, further enhancing its clinical applicability. This approach aids clinicians in early identification of high-risk patients with SAP, providing valuable support for subsequent medical decision making.

The findings indicate that patients with acute ischemic stroke complicated by SAP had a significantly poorer prognosis than those without SAP, and early identification and intervention are thus critical for improving prognosis. Using the CatBoost model, the study identifies several key factors associated SAP in patients with acute ischemic stroke including Age, NIHSS_7d, WBC_24h, SII_24h, SIRI_24h, NLR_48h, FBG, D_Dimer, G_tube, NIHSS_preMT, NIHSS_24h_postIVT. SHAP histograms ranked these eleven features in order of their influence on the model: NIHSS_7d, SIRI_24h, WBC_24h, NIHSS_preMT, D-dimer, NIHSS_24h_postIVT, G-tube, FBG, SII_24h, NLR_48h, Age.

Hyperglycemia can lead to functional defects in pulmonary dendritic cells, reduce lung immunity, and cause pulmonary immune dysfunction, thereby increasing the risk of infection(26). High NIHSS scores indicate severe neurological deficits in patients with pronounced limb paralysis and prolonged bed rest, making them susceptible to pulmonary infections and increasing the incidence of stroke-associated pneumonia. Patients who underwent intravenous thrombolysis followed by MT at our hospital were all transferred to the intensive care unit postoperatively. We observed more patients with gastric tube placement in the SAP group, indicating that gastric tube placement may increase susceptibility to SAP by increasing the risk of gastroesophageal reflux and aspiration(27). WBC is a critical marker of systemic inflammation. A significant increase in WBC suggests the presence of severe infection and systemic inflammatory response syndrome (SIRS). Age-related multisystem decline induces immunosuppression, which, combined with stroke-induced injury and aspiration risk, increases the susceptibility to stroke-associated pneumonia (SAP)(28). Elevated D-dimer levels mediate post-stroke coagulation-fibrinolysis system disorders, exacerbate vascular endothelial injury and systemic inflammatory responses, and amplify the risk of SAP in patients undergoing thrombolysis combined with thrombectomy(29).

The NLR, PLR, SII, and SIRI measure neutrophils, lymphocytes, platelets, and monocytes, reflecting various aspects of the body’s inflammatory status, immune function, and coagulation function(24). This study employed a dynamic evaluation of biomarkers at three timepoints to assess changes in inflammation and immune responses, improving the accuracy and reliability of prediction.

An elevated NLR indicates an enhanced inflammatory response and weakened immune regulation(10). Neutrophils play key roles in infection and inflammation, and their increased numbers usually reflect a considerable inflammatory response to pathogens. Lymphocytes are involved in immune regulation, and a decrease in their number may indicate suppressed immune function, creating an imbalance that may make patients more susceptible to pneumonia(30). A higher NLR in patients with acute ischemic stroke was associated with SAP and correlated with the severity of pneumonia, significantly affecting both short- and long-term patient outcomes(31). In patients with acute ischemic stroke undergoing EVT, an increased NLR served as an independent predictor of SAI, and ROC curve analysis demonstrated certain diagnostic capabilities(32). The SIRI integrates information from neutrophils, monocytes, and lymphocytes, providing a more comprehensive reflection of the systemic immune-inflammatory status. Monocyte activation releases inflammatory mediators and pro-inflammatory cytokines, exacerbating the inflammatory response. Monocytes can phagocytose pathogens; however, they may be unable to eliminate them effectively in cases of immune dysfunction, thereby increasing the risk of pneumonia(33). The SIRI has a predictive value for the occurrence of SAP in patients with acute ischemic stroke as well as for their prognosis at 3 months (34, 35). SII may be a risk factor for pneumonia in patients with acute ischemic stroke undergoing MT, highlighting the important role of platelets in inflammation and immune responses. Reperfusion therapy can lead to endothelial damage, resulting in increased platelet count and aggregation, triggering platelet activation and the release of inflammatory mediators that promote inflammatory responses. Concurrently, a reduction in lymphocytes reflects the suppression of immune function(33, 36). PLR reflects platelet aggregation and systemic inflammation (37). Previous studies have reported that PLR levels are higher in SAP patients than in non-SAP patients (38–40); however, our study did not observe a significant difference in PLR between the two groups, which may be attributed to the specific population (patients undergoing bridging therapy) or differences in sample size and baseline characteristics

ML models offer significant advantages in identifying complex patterns within clinical data, improving prediction accuracy and practical utility(41). In this study, we selected 10 common ML algorithms (including RF, LR, GBM, etc.) to construct predictive models for SAP, and compared their performance using ROC and DCA curves, leading to the selection of the CatBoost model for further analysis due to its optimal performance. The CatBoost model excels in several areas(42): (1) Automatic robust categorical feature processing; (2) Strong anti-overfitting with high generalization; and (3) Fast training and low-latency inference. Previous studies have not investigated the prediction of SAP in patients with acute ischemic stroke using multiple machine learning models, and often lacked model interpretability.

In contrast, our study is the first to construct an interpretable ML model for SAP in patients with acute ischemic stroke. We applied LASSO regression for feature selection and incorporated SHAP values to interpret the CatBoost model, thereby enhancing interpretability. SHAP summary plots revealed that eleven key variables exhibited a “bipolar separation” pattern in SHAP values, indicating their strong influence on the predicted risk of SAP.

In terms of predictive performance, the CatBoost model achieved an AUC of 0.952 in the training set. This high-performance, interpretable CatBoost model advances the risk stratification of SAP in acute ischemic stroke patients undergoing bridging therapy, facilitating early identification of high-risk individuals. Early high-risk identification enables personalized SAP prevention, and SHAP-based interpretability boosts clinical confidence and shared decision-making.

One of the advantages of this study was the identification of optimal timepoints for inflammatory markers and potential therapeutic targets. The second advantage is the application of machine learning models for SAP prediction. This study is a single-center, small-sample investigation that requires external data validation, and future large-sample, multicenter clinical trials are needed for confirmation.

## 4 Conclusion

The 24-h NLR, 24-h SII, 24-h SIRI, 48-h NLR, and 48-h SIRI were significantly associated with the occurrence of SAP in patients undergoing bridging MT after intravenous thrombolysis. The CatBoost model constructed with variables selected via LASSO regression demonstrated high predictive value for SAP, with an AUC of 0.952 in the training set.

## 6 Author Contributions

H-MW and J-TZ were responsible for the study concept and design. X-YW, M-ML, S-MZ, X-YJ, W-SY and L-LC performed the acquisition, analysis, or interpretation of data. X-YW drafted the manuscript. H-MW, J-TZ and W-SY critically revised the manuscript. M-ML and X-YW performed the statistical analyses. H-MW and J-TZ were responsible for administrative, technical, or material support.

## 8 List of Abbreviations

A2DS2: Adapted Symptom Distress Scale
AIS: Acute Ischemic Stroke
AIS-APS: Acute Ischemic Stroke-Associated Pneumonia Score
AUC: Area Under the Receiver Operating Characteristic Curve
CI: Confidence Interval
DCA: Decision Curve Analysis
EVT: Endovascular Therapy
FBG: Fasting Blood Glucose
GBM: Gradient Boosting Machine
KNN: K-Nearest Neighbors
LR: Logistic Regression
LASSO: Least Absolute Shrinkage and Selection Operator
ML: Machine Learning
MT: Mechanical Thrombectomy
mRS: Modified Rankin Scale
NIHSS: National Institutes of Health Stroke Scale
NLR: Neutrophil-to-Lymphocyte Ratio
NN: Neural Network
OR: Odds Ratio
PLR: Platelet-to-Lymphocyte Ratio
RF: Random Forest
ROC: Receiver Operating Characteristic
SAP: Stroke-Associated Pneumonia
SAIs: Stroke-Associated Infections
SII: Systemic Immune-Inflammation Index
SIRI: Systemic Inflammatory Response Index
SIRS: Systemic Inflammatory Response Syndrome
SVM: Support Vector Machine
VIF: Variance Inflation Factor
XGBoost: Extreme Gradient Boosting
LightGBM: Light Gradient Boosting Machine
AdaBoost: Adaptive Boosting
CatBoost: Category Boosting

## 9 Acknowledgments

We would like to thank Editage (https://www.editage.cn) for English language editing.

## 10 Reference styles

Vancouver (numbered)

## 11 Data Availability Statement

The raw data supporting the conclusions of this article will be made available by the authors, without undue reservation.

---

**Supplemental Figure 1.**
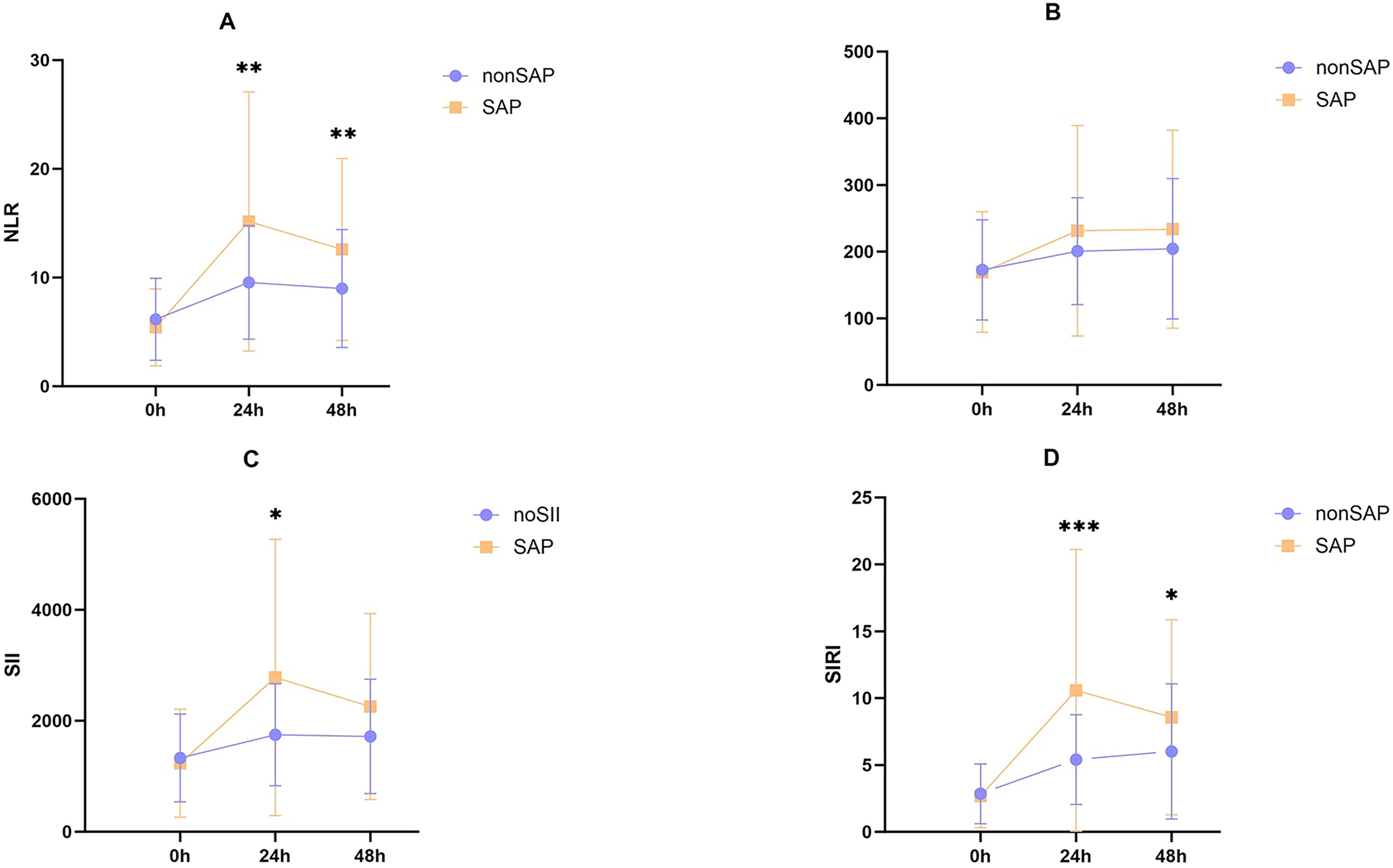
Longitudinal inflammatory marker levels in SAP and non-SAP patients.

**Supplemental Figure 2.**
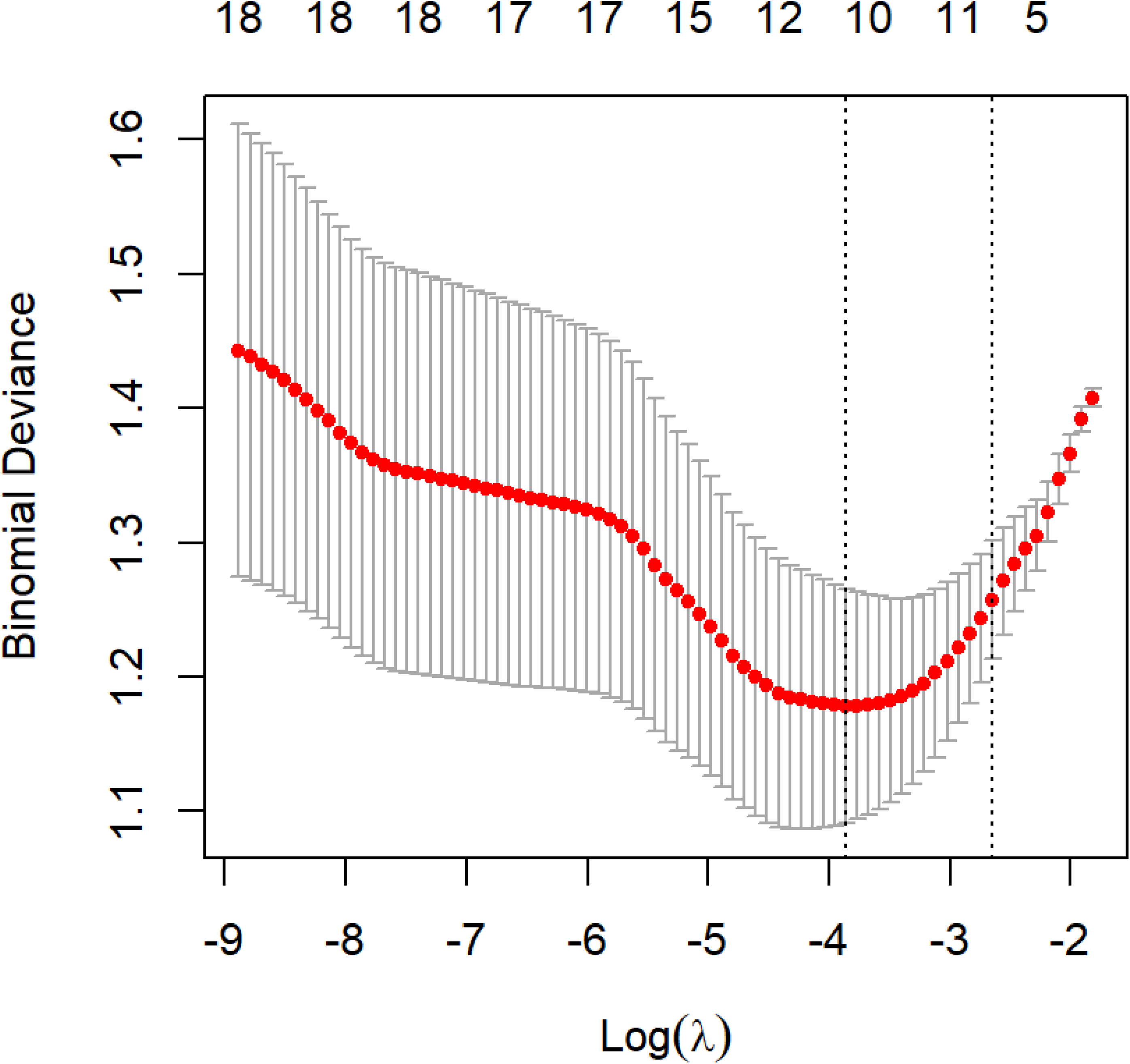
LASSO regression tuning plot for feature selection.

**Supplemental Figure 3.**
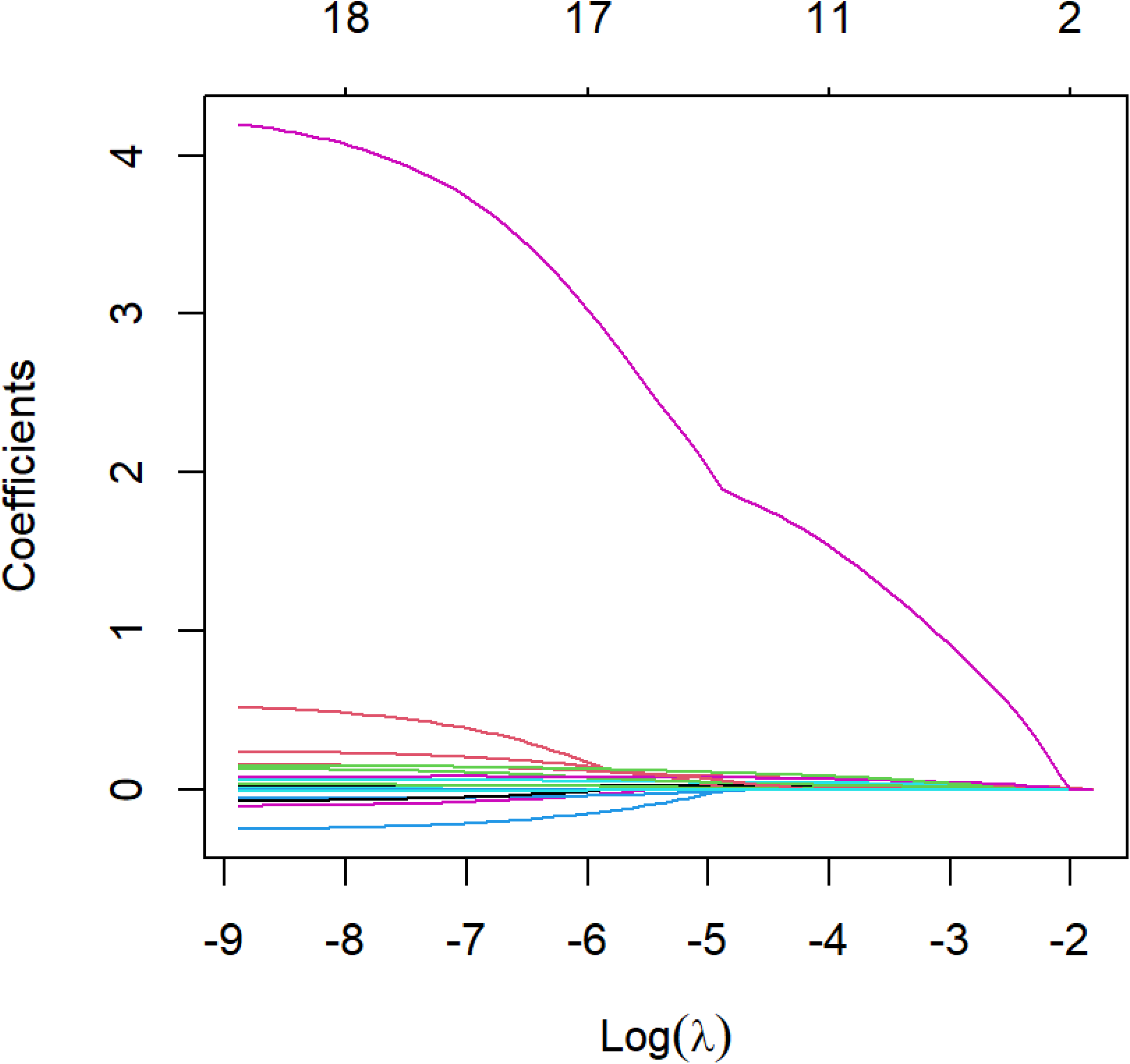
LASSO coefficient profile plot for feature selection.

**Supplemental Figure 4.**
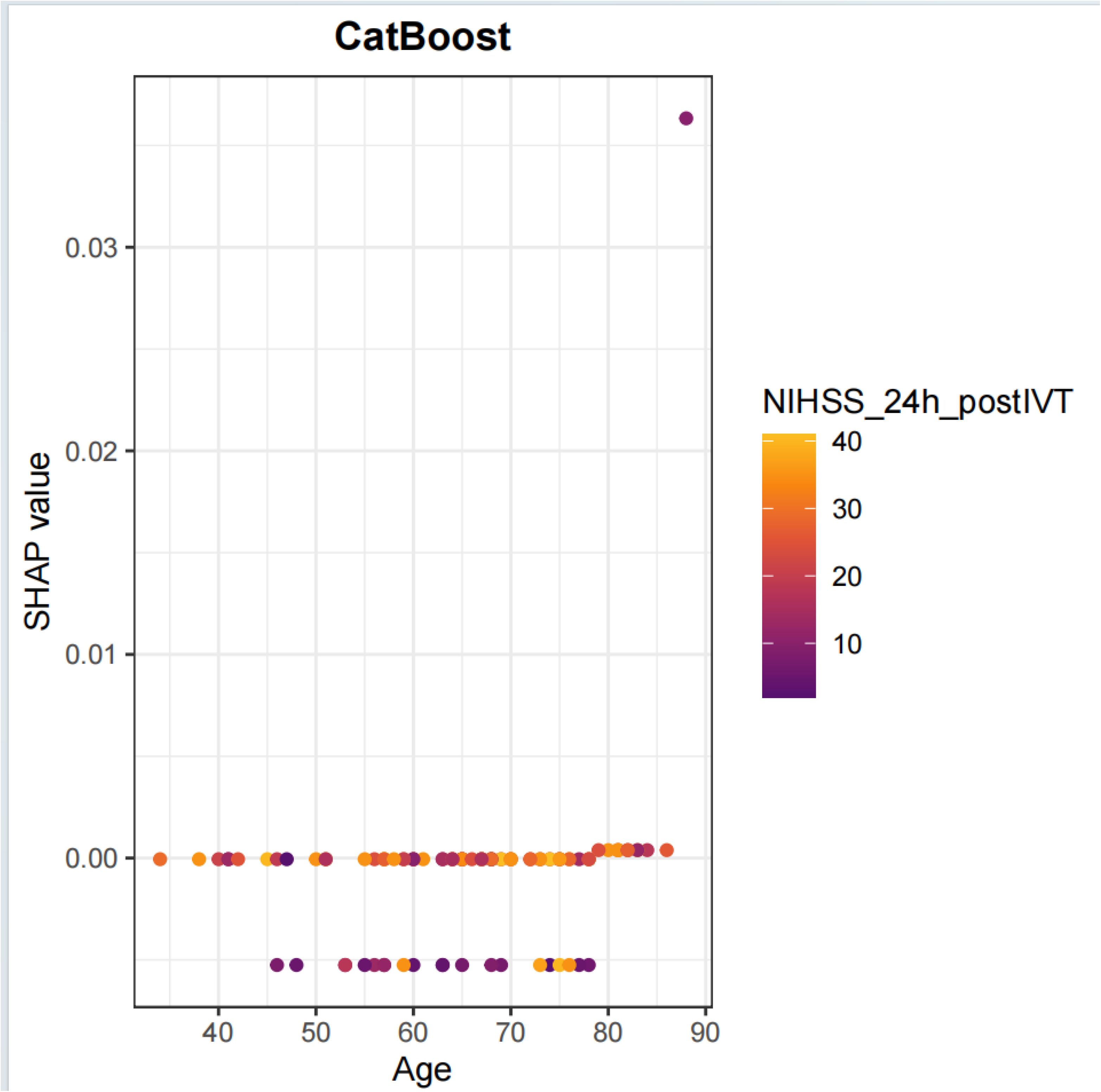
SHAP dependence plot of age for the CatBoost pneumonia prediction model.

**Supplemental Figure 5.**
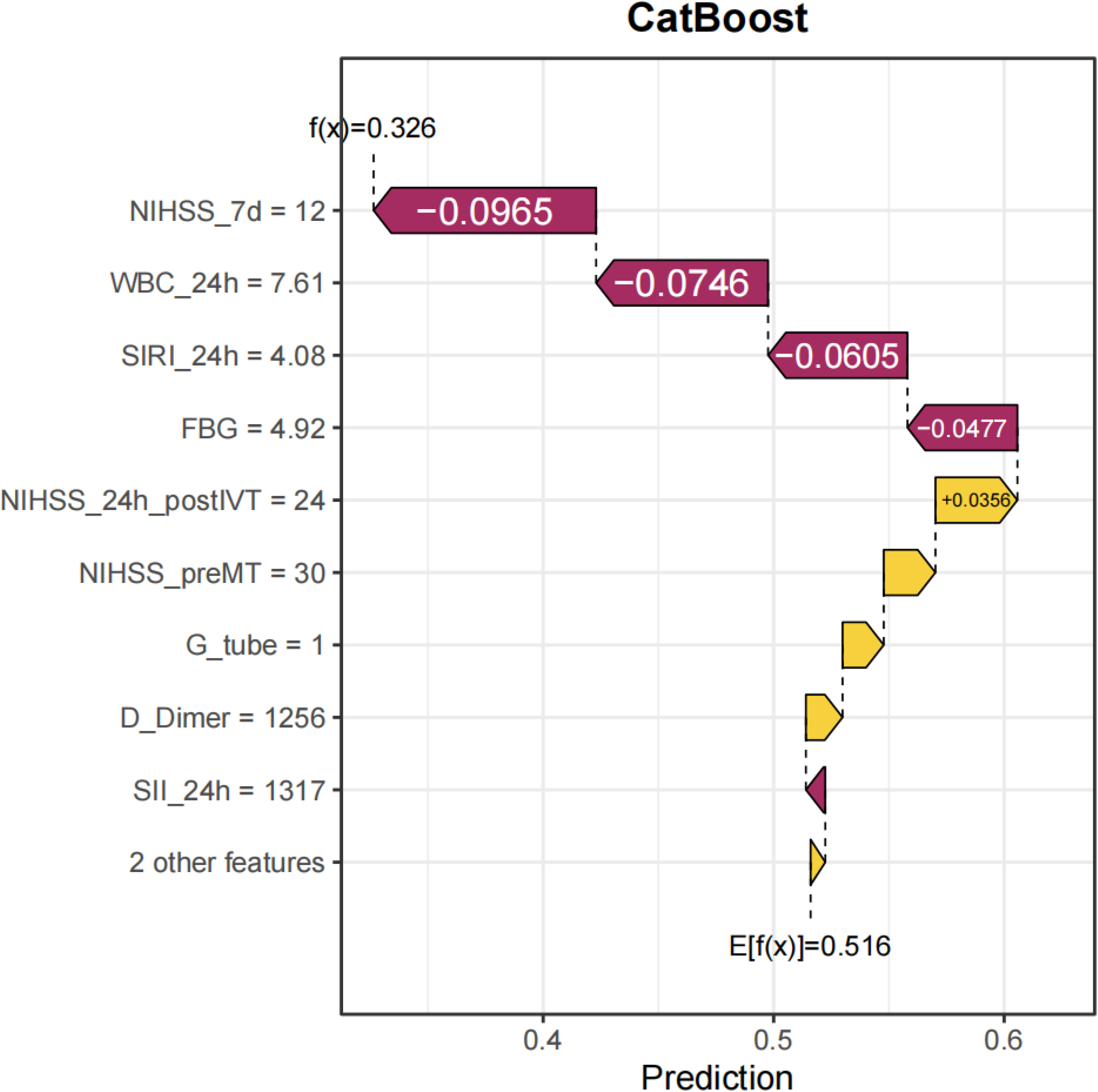
SHAP waterfall plot of the CatBoost pneumonia prediction model.

**Supplemental Figure 6.**
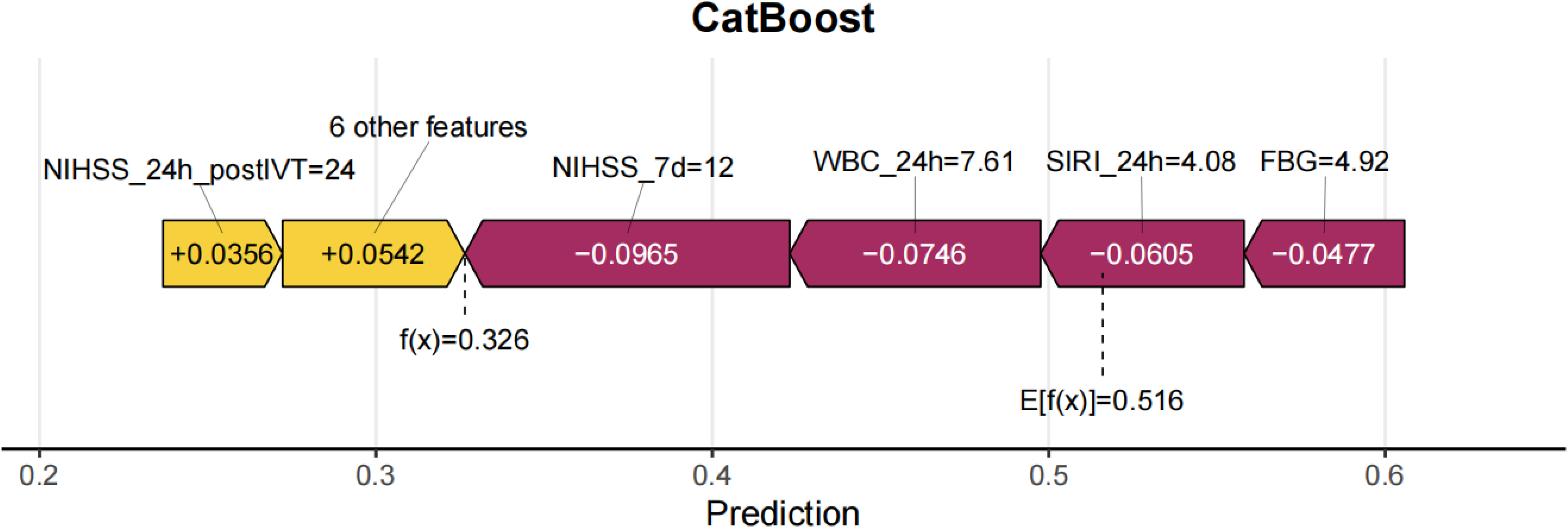
SHAP force plot of the CatBoost pneumonia prediction model.

